## Supplemental Tables for "Examining the Impact of ICU Population Interaction Structure on Modeled Colonization Dynamics of *Staphylococcus aureus*"

### Population Structure Drives Differential Methicillin-resistant *Staphylococcus aureus* Colonization Dynamics

#### Supplemental Material

**Table S1** Transitions and Equations for the Single Staff Type (SST) Model of MRSA Acquisition

| Transition | Equation |
|---|---|
| $S_C$ to $S_U$ | $\iota S_C$ |
| $S_C$ to $S_U$ | $\tau S_C \frac{P_C}{P_C + P_U}$ |
| $S_U$ to $S_C$ | $\rho \sigma S_U \frac{P_C}{(P_C + P_U)}$ |
| $P_U$ to $P_C$ | $\rho \psi P_U \frac{S_C}{(S_C + S_U)}$ |
| $P_U$ Discharge to $P_U$ Admission | $\theta \nu_U P_U$ |
| $P_U$ Discharge to $P_C$ Admission | $\theta \nu_C P_U$ |
| $P_C$ Discharge to $P_U$ Admission | $\theta \nu_U P_C$ |
| $P_C$ Discharge to $P_C$ Admission | $\theta \nu_C P_C$ |
| $P_C$ to $P_U$ | $\mu P_C$ |

**Table S2** Transitions and Equations for the Nurse-MD Model of MRSA Acquisition

| Transition | Equation |
|---|---|
| $N_C$ to $N_U$ | $\iota_N N_C$ |
| $N_C$ to $N_U$ | $\tau_N N_C \frac{P_C}{(P_C + P_U)}$ |
| $N_U$ to $N_C$ | $\rho_N \sigma N_U \frac{P_C}{(P_C + P_U)}$ |
| $D_C$ to $D_U$ | $\iota_D D_C$ |
| $D_C$ to $D_U$ | $\tau_D D_C \frac{P_C}{(P_C + P_U)}$ |
| $D_U$ to $D_C$ | $\rho_D \sigma D_U \frac{P_C}{(P_C + P_U)}$ |
| $P_U$ to $P_C$ | $\rho_N \psi P_U \frac{N_C}{(N_C + N_U)}$ |
| $P_U$ to $P_C$ | $\rho_D \psi P_U \frac{D_C}{(D_C + D_U)}$ |
| $P_U$ Discharge to $P_U$ Admission | $\theta \nu_U P_U$ |
| $P_U$ Discharge to $P_C$ Admission | $\theta \nu_C P_U$ |
| $P_C$ Discharge to $P_U$ Admission | $\theta \nu_U P_C$ |
| $P_C$ Discharge to $P_C$ Admission | $\theta \nu_C P_C$ |
| $P_C$ to $P_U$ | $\mu P_C$ |

**Table S3** Transitions and Equations for the Metapopulation Model of MRSA Acquisition

| Transition | Equation |
|---|---|
| $N_{C_i}$ to $N_{U_i}$ | $\iota_N N_{Ci}$ ; $i = 1 \dots 6$ |
| $N_{C_i}$ to $N_{U_i}$ | $\tau_N N_{Ci} \frac{P_{Ci}}{(P_{Ci} + P_{Ui})} \gamma$ ; $i = 1 \dots 6$ |
| $N_{C_i}$ to $N_{U_i}$ | $\tau_N N_{Ci} \frac{P_{Cj}}{(P_{Cj} + P_{Uj})} [(1-\gamma)/5]$ ; $i = 1 \dots 6, j = 1 \dots 6, j \neq i$ |
| $N_{U_i}$ to $N_{C_i}$ | $\rho_N \sigma N_{Ui} \frac{P_{Ci}}{(P_{Ci} + P_{Ui})} \gamma$ ; $i = 1 \dots 6$ |
| $N_{U_i}$ to $N_{C_i}$ | $\rho_N \sigma N_{Ui} \frac{P_{Cj}}{(P_{Cj} + P_{Uj})} [(1-\gamma)/5]$ ; $i = 1 \dots 6, j = 1 \dots 6, j \neq i$ |
| $D_C$ to $D_U$ | $\iota_D D_C$ |
| $D_C$ to $D_U$ | $\tau_D D_C \frac{\sum_{i=1}^{6} P_{Ci}}{\sum_{i=1}^{6} (P_{Ci} + P_{Ui})}$ |
| $D_U$ to $D_C$ | $\rho_D \sigma D_U \frac{\sum_{i=1}^{6} P_{Ci}}{\sum_{i=1}^{6} (P_{Ci} + P_{Ui})}$ |
| $P_{U_i}$ to $P_{C_i}$ | $\rho_N \psi P_{Ui} \frac{N_{Ci}}{(N_{Ci} + N_{Ui})} \gamma$ ; $i = 1 \dots 6$ |
| $P_{U_i}$ to $P_{C_i}$ | $\rho_N \psi P_{Ui} \frac{N_{Cj}}{(N_{Cj} + N_{Uj})} [(1-\gamma)/5]$ ; $i = 1 \dots 6, j = 1 \dots 6, j \neq i$ |
| $P_{U_i}$ to $P_{C_i}$ | $\rho_D \psi P_{Ui} \frac{D_C}{(D_C + D_U)}$ ; $i = 1 \dots 6$ |
| $P_{U_i}$ Discharge to $P_{U_i}$ Admission | $\theta \nu_U P_{Ui}$ ; $i = 1 \dots 6$ |
| $P_{U_i}$ Discharge to $P_{C_i}$ Admission | $\theta \nu_C P_{Ui}$ ; $i = 1 \dots 6$ |
| $P_{C_i}$ Discharge to $P_{U_i}$ Admission | $\theta \nu_U P_{Ci}$ ; $i = 1 \dots 6$ |
| $P_{C_i}$ Discharge to $P_{C_i}$ Admission | $\theta \nu_C P_{Ci}$ ; $i = 1 \dots 6$ |
| $P_{C_i}$ to $P_{U_i}$ | $\mu P_{Ci}$ ; $i = 1 \dots 6$ |
